# OrthoFx Rescue Aligner: A Mid-Course Correction during Orthodontic Clear Aligner Therapy

**DOI:** 10.1101/2023.05.29.23288751

**Authors:** Evan Chwa, Tamara Alqazaha, Flavio José Castelli Sanchez, Budi Kusnoto, Mohammed H. Elnagar

**Author notes:** Corresponding Author Mohammed H. Elnagar, DDS, MS, PhD, Department of Orthodontics, University of Illinois at Chicago, 801 S. Paulina Street, Chicago, IL 60612.

## Abstract

Over the last few decades, there has been a significant increase in the number of patients seeking orthodontic treatment due to the esthetics of clear aligners. However, clear aligners rely on accurate tracking and if not achieved, can lead to frustrations.

Data from 60 comprehensive clear aligner patients were analyzed. The OrthoFX Rescue Aligner intervened in the cases that were not tracking to get the patient back on track. The Rescue Aligner is designed to quickly and cost-effectively correct treatment lag up to a four to eight weeks deviation. The hyper elastic rescue aligners could produce movement ranges up to at least 0.5mm per tray.

This retrospective study indicates the potential for further prospective clinical and laboratory studies with improved methods such as 3D superimposition, remote monitoring as well as calibrated inter and intra observer visual inspection methods for better assessment on tracking of each tooth per aligner.

## Introduction

It is undeniable that orthodontic treatment has experienced a remarkable surge in popularity among adults over the past decades. The advent of clear aligners has only further contributed to this growth, as they provide a more aesthetically pleasing option for treatment (Lingenbrink 2002). Ultimately, clear aligners can provide a viable orthodontic solution, but there are limitations to consider. One of the key limitations is the high percentage of clear aligner cases that go off track. When this happens, backtracking or rescanning can add as much as five to ten weeks to the treatment timeline, which can be a major source of frustration for patients and the providers. This limitation significantly impedes the attractiveness of clear aligners as an appealing orthodontic solution.

Many factors can cause aligner treatments to go off-track such as patient compliance, lag (air gap between tooth and plastic), poor treatment planning, diagnosis, and viscoelastic polymer material limitation. Patient compliance is important, as aligners must be worn for 22 hours a day to be effective. Poor treatment planning and diagnosis can lead to insufficient forces required to move teeth. Finally, the plastic materials used must be thick enough to move teeth but thin enough to fit the mouth comfortably without getting caught in undercuts (Bowman 2017 and Kravitz 2009). Finding a balance between these factors is key to successful alignment.

A recent prospective clinical study found that the accuracy of the tooth movements predicted by the digital planning software was only 50%, indicating that the proposed tooth movements do not accurately reflect the reality of the movement when using clear aligners (Smith 2022). Despite initially successful treatment outcomes, the accumulation of errors in the adaptation of the polymer to the teeth may lead to compromised biomechanics of the aligner (Chisari 2014). This can cause unwanted or inefficient tooth movements and ultimately result in an unsatisfactory treatment outcome, thus unsatisfied patients.

This study looked at the data gathered from 60 clear aligner cases of which 49 were “rescued” with the OrthoFX Rescue Aligner. The rescue aligners are a Hyper Elastic branded product designed to quickly and cost-effectively correct misalignment of a single tooth or set of teeth. The aligner must fit the current dentition and at least +/- 4 stages. It provides the necessary forces to move the dentition back on track, even if there has been 4 to 8 weeks of incorrect movements.

In the past few years, thermoplastic clear aligner materials have seen significant developments and enhancements, such as multilayers for increased flexibility and durability. Popular brands of aligner materials include Invisalign’s SmartTrack, Zendura FLX, and GT FLEX. All of these materials can be categorized as thermoplastic and viscoelastic consisting of a multilayered design and combination of polymers. Invisalign’s SmartTrack is a multilayer aromatic thermoplastic polyurethane/copolyester that claims to provide faster, effective, and predictable movements (Morton 2017).

Thermoplastic polymer materials have been the go-to-choice for aligner therapy, but their intrinsic tendency to rapidly lose force over time has prompted OrthoFX to develop a new aligner that combines the thermoplasticity of current aligner materials with the hyper elasticity of rubbers, polymers, foams and sponges, and even some biological tissues.

Viscoelasticity means that a material’s elasticity is affected by the rate of deformation and the time the material is deformed (Lee 2022). They are unique in that they have both solid and liquid properties. This is demonstrated by the energy released during deformation, known as hysteresis, which is displayed by the area between the loading and unloading curve on a stress-strain plot. Other unique properties of these materials are stress relaxation, which is the decrease in stress while strain remains steady, and creep, which is the increase in strain while stress is maintained. Hyper Elastic materials can undergo very large shape changes (up to 700% strain) without losing their volume, and they can recover their original shape after the load is removed (Shahzad 2015). This makes them an ideal material for aligner therapy, as they can provide the same level of force over a longer period.

Furthermore, viscoelastic thermoplastic has a steep force curve or stress/strain relationship as shown by the black line in Figure 3. The rate of stress changes with respect to varying strains is exponential. This means that the tooth needs to only move slightly or the strain changes very little while the forces will take a very large steep decline. A Hyper Elastic material on the other hand has stress/strain relationships that are independent (the green line). Therefore, as the tooth moves, the same optimal forces required for orthodontic tooth movements are still available and have not depleted to near nothing.

Another factor that is a major cause of standard industry aligner failure today is the lack of fit flexibility. The exponential stress/strain relationship and the low elongation to yield that are intrinsic with thermoplastic aligner polymer negatively impact the fit flexibility of aligners. A hyper elastic polymer having a decoupled stress/strain relationship and a half order magnitude increase in elongation to yield, significantly increases the fit range of aligners fabricated from a Hyper Elastic polymer.

## Material and Methods

This retrospective study evaluated the effectiveness of OrthoFX rescue aligners as a treatment for when clear aligner therapy goes off track. Data from 60 de-identified patients who had previously undergone comprehensive clear aligner therapy was collected and analyzed. The study protocol was approved by WCG IRB (Study Number: 1299711, IRB Number: 20204609) Providers visually assessed the fit of the aligners and prescribed the OrthoFX Rescue Aligner when teeth deviated from the intended treatment plan. In addition to data visually, a limited laboratory test was conducted using 3D printed models derived from a few of the patients. Simulated movements were then fabricated and used to test the fit of the rescue aligners made of different materials (Figure 1). OrthoFX Hyper Elastic aligners could produce movement ranges up to at least 0.5mm per tray, thus this preliminary study tested the feasibility of fitting aligners with a range of tooth movement up to at least 0.5mm per tray. Data collected from the maxillary and mandibular arches included the current stage of treatment and the complexity of the case.

**Figure 1:**
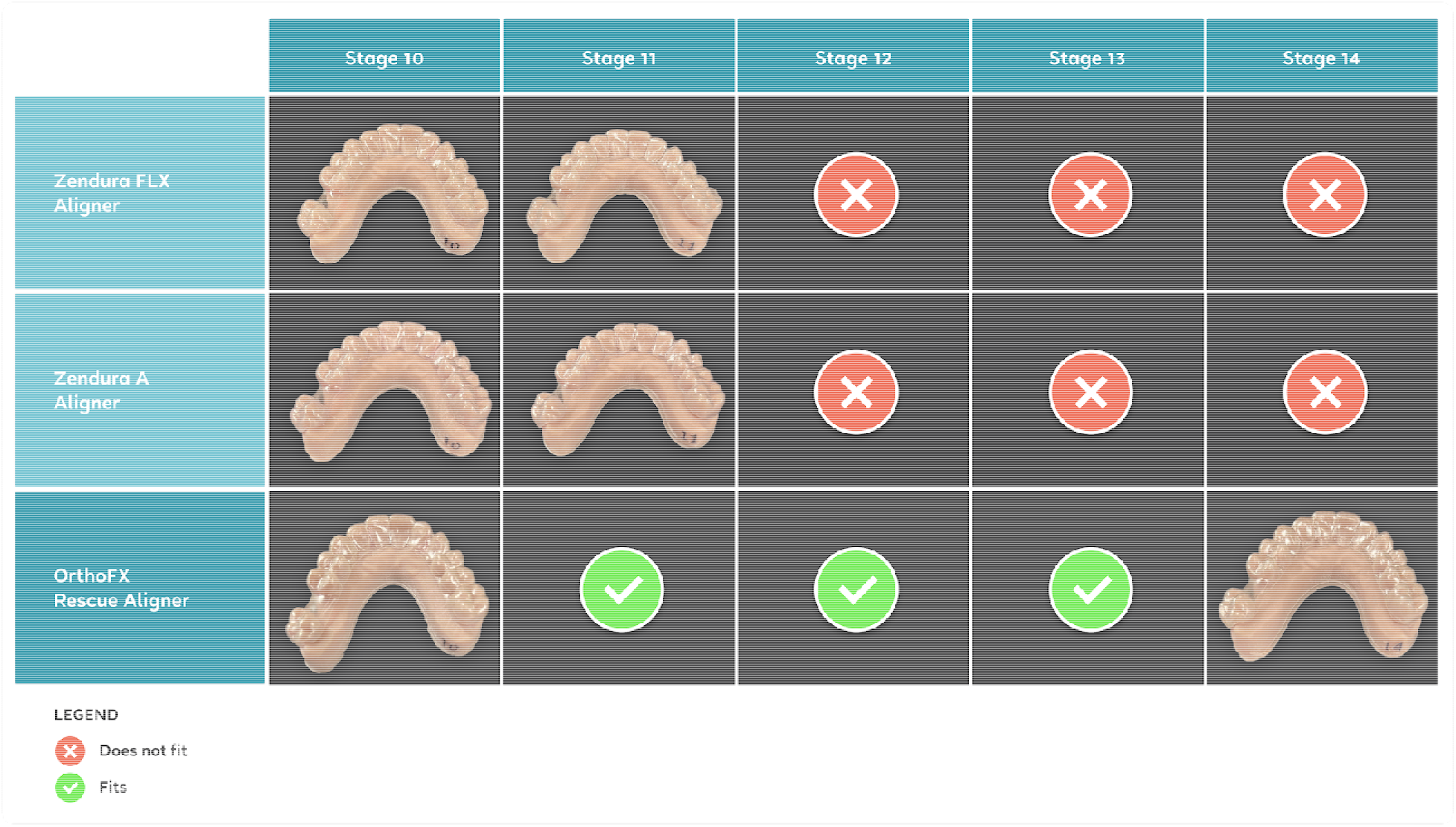
Comparison of Fitness of different materials through several stages of active aligner treatment.

The OrthoFX’s Rescue Aligner is composed of A-B-A block copolymers constructed using methods that in actual use transfer the stress energy to the strain property. The stress energy being transferred to the strain property during the hysteresis process provides greater flexibility and a greater working range. The patented OrthoFX Rescue aligner material exhibits decoupled stress and strain, hyper elastic properties, precise orthodontic movement, continuous and constant optimal forces, greater effective fitting ranges, enable the ability to produce movement span of at least +/- 0.5mm and four times more resilient force level when compared with some of the industry standard thermoplastic material. The Rescue aligner is characterized by a flat stress/strain curve, like NiTi alloy (Figure 2).

**Figure 2.**
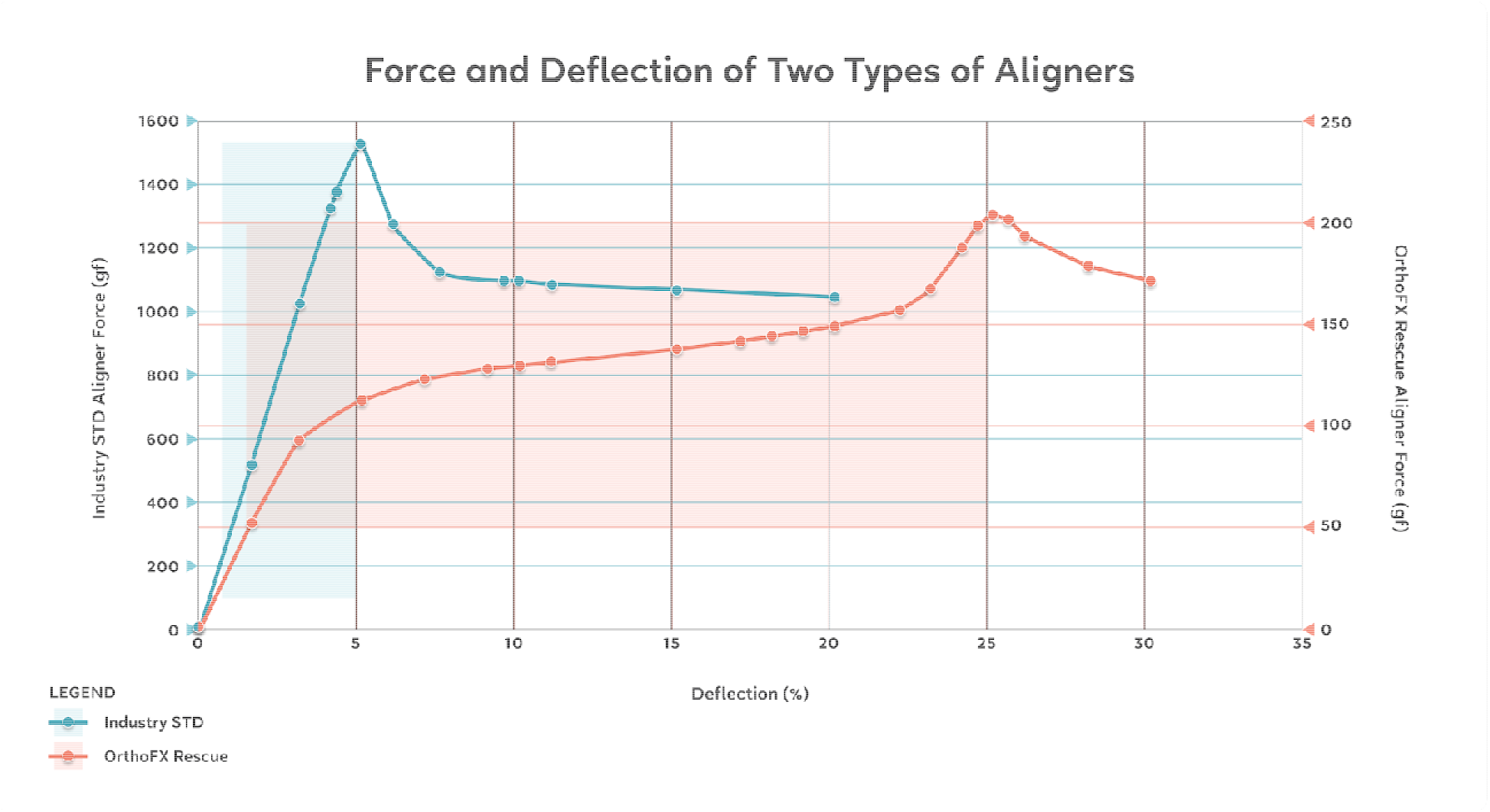
Material deflection rate comparison between OrthoFx Rescue Aligner and other standard aligners

## Results

48 of the 60 off-track subjects were successfully rescued and able to continue with the original treatment due to the Rescue Aligner. The following results pertain to the maxillary arch only. For the simple cases, 11 patients had comprehensive treatment requiring less than 17 trays and the Rescue Aligner was used approximately 63% ± 24% into treatment. For the moderate cases, 30 had comprehensive treatment requiring 17-34 trays and the Rescue Aligner was used 50% ± 17% into treatment. For the complex subjects, 9 had comprehensive treatment requiring over 34 trays and the Rescue Aligner was used on average 47% ± 21% into treatment. In general, Rescue Aligners were used midway in treatment being 30-70% with a Pearson’s correlation of 0.487. For the mandibular arch, only 11 of the 60 subjects untracked who needed to be corrected via the wear of Rescue Aligners. Of the cases, the Rescue Aligner was used 62% ± 22% into treatment. In total, an average of 25 ± 9 stages were documented with the aligner prescribed 53% ± 19% into treatment.

**Table 1:** Percentage Utilization of Rescue Aligners from total number of aligners prescribed during active treatment

| MAXILLARY | Number of subjects | Average Percentage of number of trays into treatment when rescue initiated | SD |
| --- | --- | --- | --- |
| Mild | 11 | 63% | 24% |
| Moderate | 40 | 51% | 17% |
| Severe | 9 | 47% | 21% |

| MANDIBULAR | Number of subjects | Average Percentage of number of trays into treatment when rescue initiated | SD |
| --- | --- | --- | --- |
| Mild | 1 | 1% | 4% |
| Moderate | 9 | 3% | 8% |
| Severe | 1 | 4% | 14% |

## Discussion

The OrthoFX Rescue aligner combines the benefits of thermoplastic and Hyper Elastic materials, making it significantly more efficient and effective than other thermoplastic and viscoelastic aligner materials

Most orthodontic movements can be achieved with just 50 grams of force. However, an industry standard aligner will exert 7.5 grams orders of magnitude of force greater than the minimum force to begin orthodontic movement. Then approximately 3% later in deflection, the force being exerted evaporates to almost nothing (Figure 2).

Results for the maxillary arch showed that the more complex the case, the earlier the Rescue aligner would be used. In milder cases, the Rescue aligner was used more at the end of treatment. The success rate of the Rescue aligner was 82%, with the maxillary lateral incisors being the main teeth that had difficulty tracking. Out of the 11 cases that couldn’t be rescued, one was simple, seven were moderate, and three were complex. The most difficult movement to correct was extrusion, followed by tipping and rotation.

It was difficult to determine exactly when the Rescue Aligner was utilized earlier in the treatment process for complex cases, while milder cases had better tractability when the aligner was applied at the end of treatment. In this sample, the incidence of untracked aligners in the lower arch needing Rescue Aligners was found to be much less than in the upper arch. The data of the lower arch (n=-.194) was still included, but with smaller sample size. It was hypothesized that subjects mainly focused on the maxillary teeth traction and not the mandibular teeth due to aesthetics. Although a smaller sample size, showed the lower arch are similar to that of the upper arch. This feasibility preliminary retrospective study shows the potential for further clinical and laboratory prospective study. Further clinical studies can observe inter and intra reliability with additional calibration for the visual inspection interpretation as well as integrating more robust methods such as 3D superimposition and remote monitoring technology for better tracking of each tooth per aligner.

## Conclusion

The concept of Rescue Aligners has not been fully explored in the field of clear aligner therapy. The specific aim of this study was to test the feasibility of using new materials of aligners to avoid midcourse corrections and refinements. In this preliminary study, the use of Rescue Aligners shows its efficacy in correcting off tracked cases. Future prospective study will include 3D superimposition and remote monitoring technology to study the efficacy of tooth movement individually.

## Data Availability

All data produced in the present work are contained in the manuscript

## Acknowledgements

All data was collected by OrthoFX. The OrthoFX Rescue aligner is FDA cleared and meets all ISO-10993 biocompatibility requirements.

